# Extremely preterm infant admissions within the SafeBoosC-III consortium during the COVID-19 lockdown

**DOI:** 10.1101/2020.10.02.20204578

**Authors:** Marie Isabel Rasmussen, Mathias Lühr Hansen, Gerhard Pichler, Eugene Dempsey, Adelina Pellicer, Afif EL-Khuffash, A Shashidhar, Salvador Piris-Borregas, Miguel Alsina, Merih Cetinkaya, Lina Chalak, Hilal Ozkan, Mariana Baserga, Jan Sirc, Hans Fuchs, Ebru Ergenekon, Luis Arruza, Amit Mathur, Martin Stocker, Olalla Otero Vaccarello, Tomasz Szczapa, Kosmas Sarafidis, Barbara Krolak-Olejnik, Asli Memisoglu, Hallvard Reigstad, Elżbieta Rafińska-Ważny, Eleftheria Hatzidaki, Zhang Peng, Despoina Gkentzi, Renaud Viellevoye, Julie De Buyst, Emmanuele Mastretta, Ping Wang, Gitte Holst Hahn, Lars Bender, Luc Cornette, Jakub Tkaczyk, Ruth del Rio, Monica Fumagalli, Evangelina Papathoma, Maria Wilinska, Gunnar Naulers, Iwona Sadowska-Krawczenko, Chantal Lecart, María Luz Couce, Siv Fredly, Anne Marie Heuchan, Tanja Karen, Gorm Greisen

**Affiliations:** Department of Neonatology, Rigshospitalet, Copenhagen, Denmark; Department of Pediatrics, Medical University of Graz, Graz, Austria; Infant Centre and Department of Paediatrics and Child Health, University College Cork, College Road, Cork, Ireland; Department of Neonatology, La Paz University Hospital, Madrid, Spain; Department of Pediatrics, The Royal College of Surgeons in Ireland, Dublin, Ireland; St. Johns Medical College Hospital, Bengaluru, India; Department of Neonatology, 12 Octubre University Hospital. Madrid; Neonatology Department, Hospital Clínic-Maternintat, Barcelona, Spain; Department of Neonatology, Kanuni Sultan Suleyman Training and Research Hospital, Küçükçekmece/İstanbul, Turkey; Division of Pediatrics - Neonatal-Perinatal, UT Southwestern, United States; Division of Neonatology, Department of Pediatrics, Uludag University Medical Faculty, Turkey; Division of Neonatology, Department of Pediatrics, University of Utah, United States; Institute for the Care of Mother and Child, Prague, Czech Republic; Center for Pediatrics, Department of Neonatology, Medical Center, University of Freiburg, Germany; Department of Neonatology, Gazi University Hospital, Yenimahalle/Ankara, Turkey; Division of Neonatology. Instituto del Niño y del Adolescente. Hospital Clinico San Carlos-IdISSC, Madrid, Spain; Department of Neonatal-Perinatal Medicine, Saint Louis University School of Medicine, Missouri, United States; Neonatal and Pediatric Intensive Care Unit, Children’s Hospital Lucerne, Switzerland; Department of Neonatology, Hospital Universitario de Tarragona Juan XXIII, Tarragona, Spain; Department of Neonatology, Neonatal Biophysical Monitoring and Cardiopulmonary Therapies Research Unit, Poznan University of Medical Sciences, Poznań, Poland; First Department of Neonatology, Aristotle University, Hippokrateion General Hospital, Thessaloniki, Greece; Department of Neonatology, Wrocław Medical University, Poland; Department of Neonatology, Marmara University Pendik Training and Research Hospital, Pendik/Istanbul, Turkey; Department of Neonatology, Haukeland University Hospital, Bergen, Norway; Department of Neonatology, Centrum Medyczne “Ujastek”, Krakow, Poland; Department of Neonatology & NICU, University Hospital of Heraklion, Crete, Greece; Department of Neonatology, Children’s Hospital of Fudan University, Shanghai, China; NICU, Department of Pediatrics, University General Hospital of Patras, Patras, Greece; Department of Pediatrics, Neonatal Intensive Care Unit, CHU – CHR Liège, Belgium; NICU, Tivoli Hospital, La Louviere, Belgium; S.C. Neonatologia - Pres Osp S.Anna – Citta della Salute e della Scienza di Torino, Italy; Department of Neonatology, Guangzhou Women and Children’s Medical Center, Guangzhou, China (?); Department of Neonatology, Aalborg Universitets Hospital, Aalborg, Denmark; Department of Neonatology, AZ St-Jan Bruges, Belgium; Department of Neonatology, University Hospital Motol, Prague, Czech Republic; Department of Neonatology, Hospital Sant Joan de Déu, Barcelona, Spain; Department of Neonatology, Fondazione IRCCS Ca’ Granda Ospedale Maggiore Policlinico Milan, Milan, Italy; Department of Clinical Sciences and Community Health, University of Milan, Italy; Neonatal Intensive Care Unit, “Alexandra” University and State Maternity Hospital, Athens, Greece; Neonatology Department, Centre of Postgraduate Medical Education, Warsaw, Poland; Department of Neonatology, University Hospital Leuven, Leuven, Belgium; Department of Neonatology, Collegium Medicum in Bydgoszcz Nicolaus Copernicus University in Toruń,, Poland; Department of Neonatology, GHdC Charleroi, Belgium; Neonatology Department, University Clinical Hospital of Santiago de Compostela, Health Research Institute of Santiago de Compostela, Spain; Department of Neonatology, Oslo University Hospital, Norway; Department of Neonatology, Royal Hospital for Children, United Kingdom; Department of Neonatology, University Hospital Zurich, Switzerland

## Abstract

**Objective:** To evaluate if the number of admitted extremely preterm (EP) infants (born before 28 weeks of gestational age) has changed in the neonatal intensive care units (NICUs) of the SafeBoosC-III consortium during the global lockdown when compared to the corresponding time period in 2019.

**Design:** This is a retrospective, observational study. Forty-six out of 79 NICUs (58%) from 17 countries participated. Principal investigators were asked to report the following information: 1) Total number of EP infant admissions to their NICU in the three months where the lockdown restrictions were most rigorous during the first phase of the COVID-19 pandemic, 2) Similar EP infant admissions in the corresponding three months of 2019, 3) the level of local restrictions during the lockdown period and 4) the local impact of the COVID-19 lockdown on the everyday life of a pregnant woman.

**Results:** There was no significant difference between the number of EP infant admissions during the three most rigorous lockdown months of the COVID-19 pandemic compared to the corresponding three months in 2019 (n=428 versus n=457 respectively, p=0.33). There were no significant changes within individual geographic regions and no significant association between the level of lockdown restrictions and change in the number of EP infant admissions (p=0.334).

**Conclusion:** This larger *ad hoc* study did not confirm previous studies’ report of a major reduction in the number of extremely preterm births during the first phase of the COVID-19 pandemic.

## Background

On the 11^th^ of March 2020, COVID-19 was declared a pandemic by the World Health Organisation, which led to an almost worldwide lockdown (1). During the lockdown, reductions in extremely preterm birth rates and extremely low birth weight infants have been reported in Danish (2) and Irish (3) studies. The Danish study reported a decrease in the number of infants born extremely preterm (EP, infant born before 28 weeks gestational age). The study reported one EP infant from the 12^th^ of March to the 14^th^ of April of 2020 compared to a mean of 11.4 over the same time period of the preceding five years, in all of Denmark (2). The same trend was seen in the Irish study, relying on data from one hospital in Ireland, where no extremely low birth weight infants (<1000gm) were born from January to April in 2020, compared to a mean of 4.9 from January to April during the preceding 20 years (3). A Nepalese study (4) based on nine health institutions reported the contrary, with an increase in the preterm birth rate (infants born before 37 weeks gestational age) from 16.7% (2125 preterm births out of 13189 births) between 1^st^ of January to 30^th^ of May 2019 to 20.0% between 1^st^ of January to 30^th^ of May 2020 (1342 preterm births out of 7165 births). However, as can be seen in the numbers, the overall registered number of births in the time period dropped from 13189 in 2019 to 7165 in 2020. Thus, it is plausible that healthy mothers with uncomplicated pregnancies delivering at term age might have given birth outside the hospitals, while babies delivered preterm may have happened at the hospitals as usual (4).

Regarding the Danish and Irish studies, the number of extremely preterm and extremely low birth weight infants were small and thus, the results should be interpreted with caution (5). An article in the New York Times additionally reported that several neonatologists from neonatal intensive care units (NICU) worldwide, had observed a decrease in local prematurity rates, while other neonatologists observed the contrary (6). The same observations were reported within the SafeBoosC-III consortium through verbal communication.

The COVID-19 pandemic has disrupted clinical trials across the world and affected the ability to conduct trials in a safe and effective manner (7). Thus, many trials have slowed down, been suspended, or even terminated, due to the difficulties of conducting research under lockdown conditions (7). The SafeBoosC-III trial investigates the benefit and harms of treatment guided by cerebral near-infrared spectroscopy monitoring compared with treatment and monitoring as usual in extremely preterm infants (8). Despite having a vulnerable population, potentially at increased risk of complications to a COVID-19 infection, the trial was able to proceed in most countries.

Only a few NICUs were forced to suspend enrolment. During the COVID-19 lockdown, NICUs proceeded with trial preparation tasks and the number of NICUs open for randomisation was steadily increasing. However, despite a 60% increase in the number of hospitals open for randomisation in the SafeBoosC-III trial from January to May, the average monthly number of randomisations per NICU dropped drastically in March, simultaneously with the spread of COVID-19 across Europe, as can be seen in figure one. Given the contradictory reports in the published studies on prematurity rates during the COVID-19 lockdown, and the variability in observations from neonatologists and investigators worldwide, we decided to examine the effect of the COVID-19 pandemic on prematurity rates further. For the SafeBoosC-III trial, the change in EP admissions is most relevant, as this is the eligibility criteria. The purpose of this study is therefore to evaluate, if the number of admitted EP infants has changed in NICUs within the SafeBoosC-III consortium during the global lockdown. Furthermore, we wish to evaluate if there is a difference within geographical regions, or an association of the level of local lockdown restrictions and change in the number of EP infant admissions.

## Study design and methods

This is a retrospective, observational study, based on the NICUs in the consortium of the SafeBoosC-III randomised clinical trial (8). The principal investigators from all of the 79 NICUs in the consortium, were invited to participate in this study by e-mail and asked to report the following information: 1) the number of EP infants admitted to their NICU within the three months, where the lockdown restrictions were most rigorous during the first phase of the COVID-19 pandemic, 2) the number of EP infants admitted within the corresponding three months of 2019, and 3) the level of restrictions imposed upon the public, during the most rigorous three months of the lockdown period, in a Likert scale format from one to five. The consecutive three months where the lockdown restrictions were most rigorous during the first phase of the COVID-19 pandemic were subjectively defined by local principal investigators. The scale used to classify the level of lockdown restriction, is a modified scale inspired from the New Zealand COVID-19 alert system (9), with one being the normal state of society, two being mild restrictions, three being moderate restrictions, four being strong restrictions and five being very strong restrictions (full scale in appendix 1). Principal investigators reported that the data on the number of EP infant admissions was collected from admissions logbooks, NICU databases, hospital databases, and in one case, from a national registry.

Investigators were also asked to categorise the impact of the COVID-19 lockdown on the everyday life of a pregnant woman, and additionally, if they believed that the lockdown restrictions could possibly lead to a non-admittance of EP infants (e.g. intrauterine death due to delayed admittance of the mother, no possibility to transfer the baby from place of birth to a tertiary centre). Lastly, investigators were asked if there within the last year had been any major changes in the organisation of perinatal care in their area/region, which may have changed the number of EP infant admissions to their respective NICU. The full data report template can be found in appendix 1.

### Outcomes

The primary outcome was the difference in the total number of EP infant admissions during the three months with the most rigorous lockdown restrictions during the first phase of the COVID-19 pandemic, compared to the corresponding three months in 2019.

Secondary outcomes were

1. The change in the number of EP infant admissions within the following regions: Asia, Eastern Europe, Southern Europe, Northern Europe, Western Europe, North America
2. The correlation between the level of the local lockdown restrictions and change in the number of EP infant admissions.

Exploratory outcomes were 1) the likelihood that restrictions inside or outside health institutions in the investigators country/region, have led to non-admittance of EP infants, 2) if any major changes in the organisation of perinatal care had occurred locally, which may have changed the number of admissions of EP infants and 3) the impact of the COVID-19 lockdown on the everyday life of a pregnant woman.

### Statistical analysis

The total number of EP infant admissions, during the three months with the most rigorous lockdown restrictions in 2020, during the corresponding three months in 2019, within each region and within each level of lockdown restriction, were reported as numbers (n). The primary outcome, as well as the secondary outcome regarding the change in the number of EP infant admissions within each geographical region, were analysed using Chi-square tests for 1×2 tables. To analyse the correlation between the local level of lockdown restrictions and the change in number of EP infant admissions, we used simple linear regression. The exploratory outcomes did not undergo statistical analysis but were reported and discussed. For the primary outcome, an alfa level of 5% was chosen as a threshold for significance. To correct for multiple testing in the secondary outcomes, we chose an alfa level of 1%. Statistics were conducted in IBM SPSS Statistics 25 (IBM, Armonk, NY, US).

### Sample size calculation

In a previous funding application for the SafeBoosC-III trial, we estimated that the 93 NICUs taking part in the application, had a total of 3000 EP infant admissions per year. Therefore, we expected that each of the participating NICUs would have approximately 30 admissions in 2019, i.e. approximately seven admissions in a three months period. Therefore, if half of the NICUs in the SafeBoosC-III consortium (i.e. 40 NICUs) participated, we would expect a total of 280 EP infant admissions within the 40 NICUs in 2019. Thus, a 16.5% change in the primary outcome would be needed, to show a statistical significance with a 5% alfa level as threshold.

Predefined regions based on the active SafeBoosC-III NICUs can be found in table 1

**Table 1:** predefined regions within the 79 NICUs in the SafeBoosC-III consortium

| Asia | Eastern Europe | Southern Europe | Northern Europe | Western Europe | North America |
| --- | --- | --- | --- | --- | --- |
| India, China | Austria, Poland, Czech Republic, Ukraine, Turkey | Spain, Portugal, Italy, Greece | Denmark, Norway | Germany, UK, Ireland, Switzerland, Belgium | USA |

### Ethical considerations

The Danish National Committee on Health Research Ethics ruled that this study did not constitute a health research project, therefore permission to conduct this study was not necessary. To our knowledge, two participating European NICUs consulted with their local ethical committee, regarding the need for approval, but this was not required either. This study was registered on clinicaltrials.gov (NCT04527601)

## Results

A total of 885 EP infants were admitted to 46 NICUs across 17 countries (Austria, Belgium, China, Czech Republic, Denmark, Germany, Greece, India, Ireland, Italy, Norway, Poland, Switzerland, Spain, Turkey, UK, USA); 428 during the three months with the most rigorous lockdown restrictions in 2020, and 457 in the corresponding three months of 2019 (p=0.33). Furthermore, no significant differences were found in the number of admissions between 2020 and 2019 within the individual six predefined geographical regions (table 2, fig. 2).

**Table 2:**
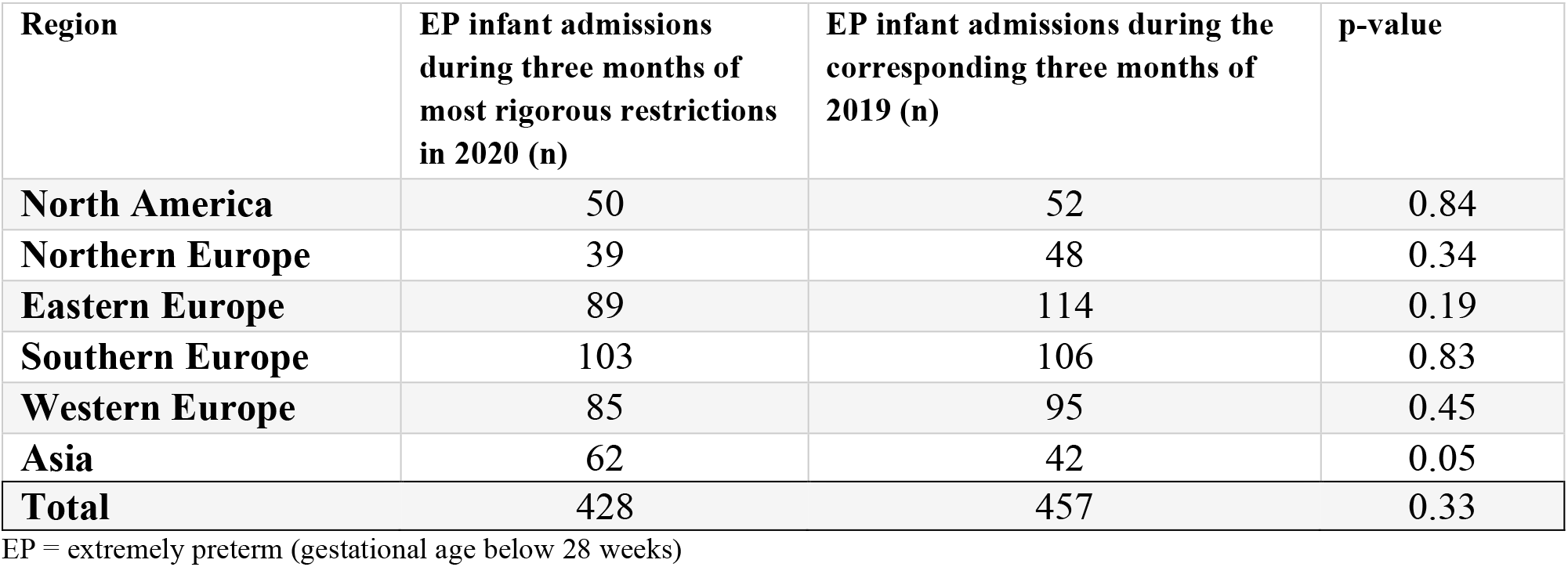
EP infant admissions in 2019 and 2020 within each predefined geographic region

| Region | EP infant admissions during three months of most rigorous restrictions in 2020 (n) | EP infant admissions during the corresponding three months of 2019 (n) | p-value |
| --- | --- | --- | --- |
| North America | 50 | 52 | 0.84 |
| Northern Europe | 39 | 48 | 0.34 |
| Eastern Europe | 89 | 114 | 0.19 |
| Southern Europe | 103 | 106 | 0.83 |
| Western Europe | 85 | 95 | 0.45 |
| Asia | 62 | 42 | 0.05 |
| **Total** | **428** | **457** | **0.33** |

EP = extremely preterm (gestational age below 28 weeks)

**Figure 1:**
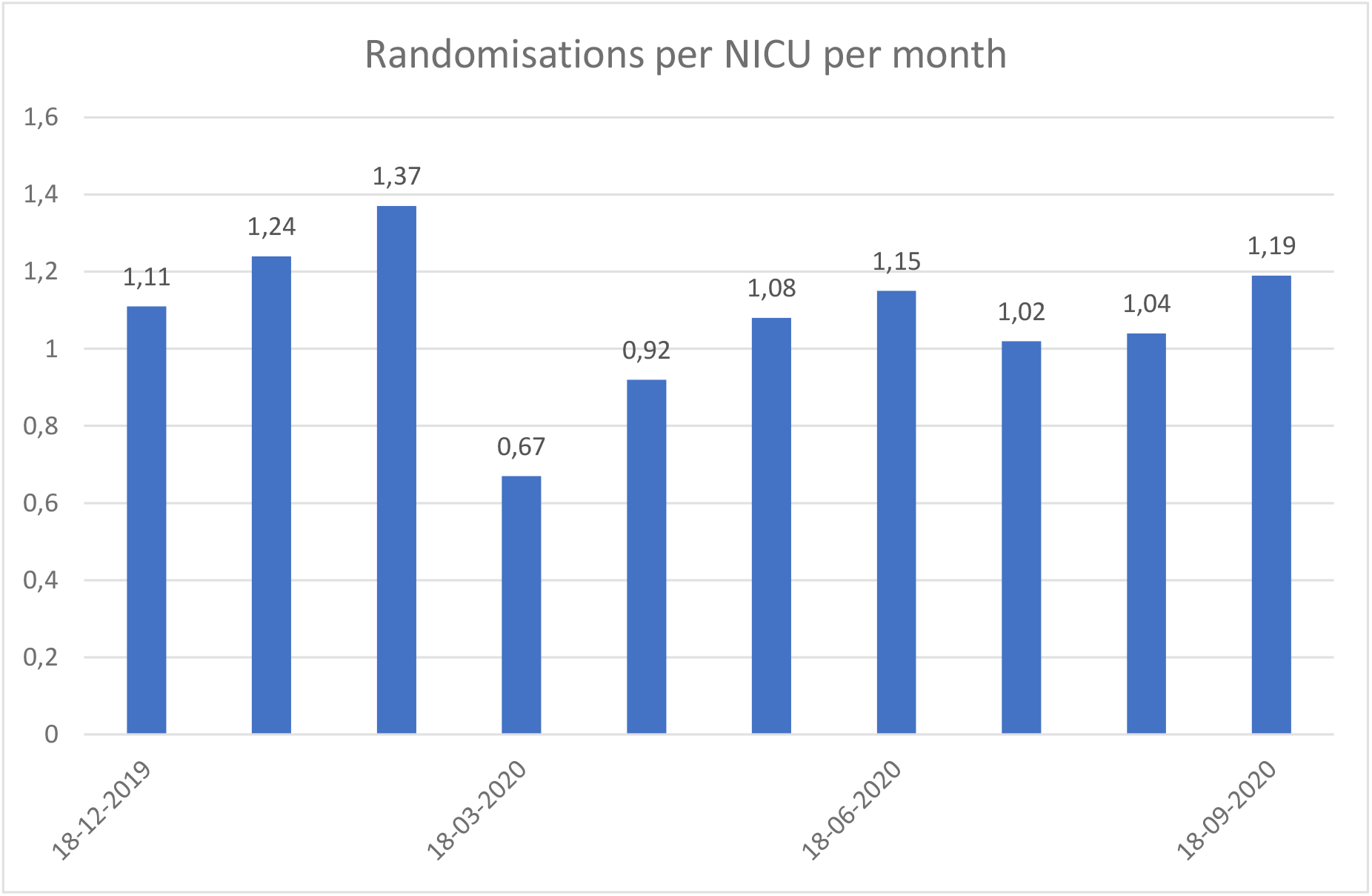
Number of randomisations per month within each NICU actively randomising infants in the SafeBoosC-III trial during the last nine months.

**Figure 2.**
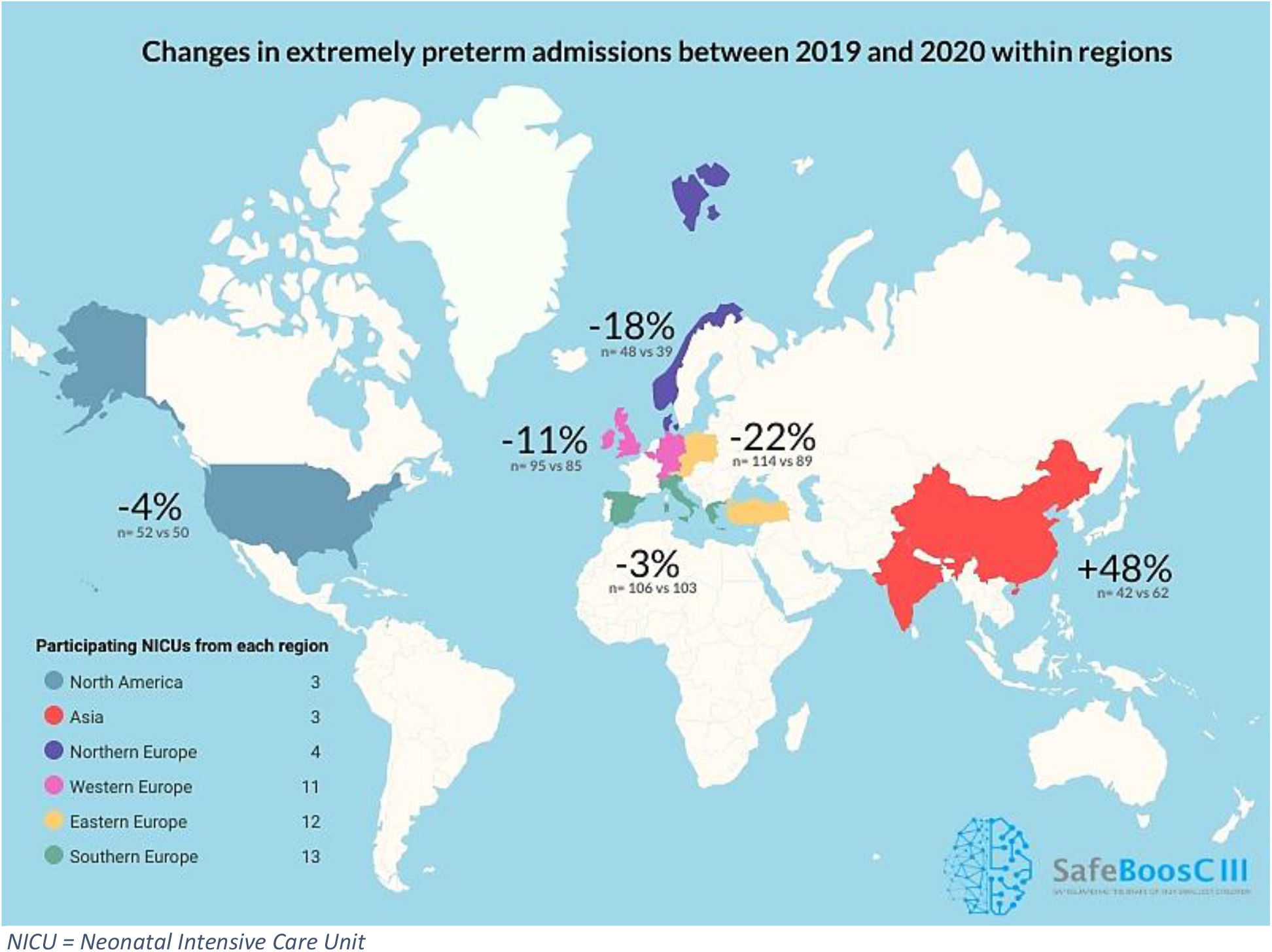
Change in percent in extremely preterm admissions between the three months with the most rigorous lockdown restrictions in 2020, compared to the corresponding months of 2019 in the 46 participating NICUs in the SafeBoosC-III consortium.

The linear regression analysis showed no significant correlation between the level of lockdown restriction and change in the number of EP infant admissions (p=0.3) (table 3).

**Table 3:** Number of EP infant admissions in 2019 and 2020 stratified by level of lockdown restriction

| Level of lockdown restrictions | EP infant admissions during the three months with the most rigorous lockdown restrictions in 2020 (n) | EP infant admissions during the corresponding three months of 2019 (n) |
| --- | --- | --- |
| 1 (no difference) | - | - |
| 2 | - | - |
| 3 | 42 | 37 |
| 4 | 64 | 66 |
| 5 (most severe) | 322 | 354 |

When investigators were asked to describe the impact of the COVID-19 lockdown on the everyday life of a pregnant woman, 33 out of 46 (72%) answered that they thought the change had been radical or very radical (table 4). Two out of 46 (4%) answered they thought the change had been little or very little. Investigators were furthermore asked to classify the likelihood that restrictions outside or inside health institutions in their country/region, could have led to non-admittance of EP infants. Thirty-four out of 46 (74%) answered that they thought it was unlikely or very unlikely (table 4). Three out of 46 (7%) answered that they thought it was likely or very likely.

**Table 4:**
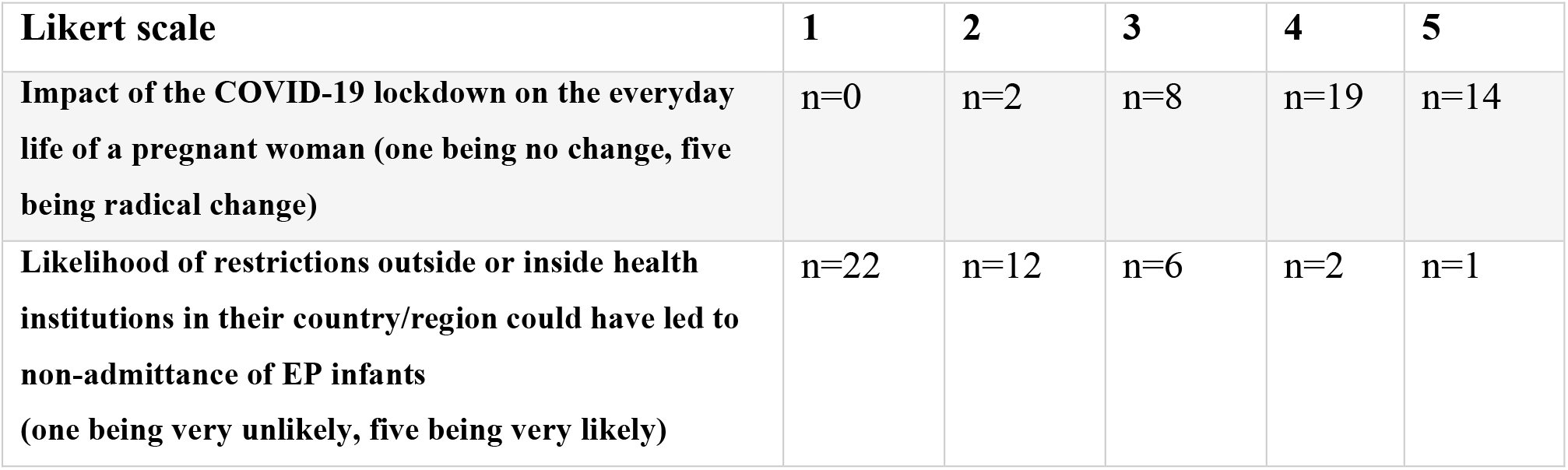
reporting of exploratory outcome results

| **Likert scale** | **1** | **2** | **3** | **4** | **5** |
| --- | --- | --- | --- | --- | --- |
| **Impact of the COVID-19 lockdown on the everyday life of a pregnant woman (one being no change, five being radical change)** | n=0 | n=2 | n=8 | n=19 | n=14 |
| **Likelihood of restrictions outside or inside health institutions in their country/region could have led to non-admittance of EP infants (one being very unlikely, five being very likely)** | n=22 | n=12 | n=6 | n=2 | n=1 |

Of the 46 participating investigators, seven reported of changes in the structure and organisation of perinatal care within their NICU. This included reports of nursing staff being rotated to COVID-19 services, increase or decrease in the NICU bed capacity due to structural changes caused by COVID-19 and COVID-19 testing of all patients before admission. One investigator also suspected that online consultations instead of on-site consultations with mothers could have led to a greater amount of intrauterine complications and thus, preterm births.

## Discussion

Based on 885 EP infant admissions from 46 NICUs across 17 countries, this international retrospective, observational study found no significant difference in the number of EP infant admissions during the three months with the most rigorous lockdown restrictions during the first phase of the COVID-19 pandemic compared with the corresponding three months in 2019. Furthermore, no significant differences in number of EP infant admissions were seen within the geographical regions. There was no correlation between levels of lockdown restrictions and change in the number of EP infant admissions.

To our knowledge, this study is the largest so far, evaluating the effect of the first phase of the COVID-19 pandemic on EP infant admissions. We exceeded the indicative sample size, which decreases the risk of type II errors, and to some extent compensates for the correction for multiple testing in the secondary outcomes (10). Furthermore, the protocol for this study was registered at clinicaltrials.gov before analysis of any data, thereby reducing the risk of selective outcome reporting bias (11).

As COVID-19 is an emergency, we decided to move quickly and therefore, the principal investigators only had three weeks (Aug 28^th^ -Sep 13^th^, 2020) to report the data. It is plausible that the participation would have been better if we had prolonged the data collection time. Thirty-three NICUs of the SafeBoosC-III consortium did not participate in this study, for reasons which are uncertain. This imposes a potential bias. Furthermore, the data was self-reported, not verified by a third party, and collected in an unblinded fashion. We compared only with the corresponding time period in 2019. Numbers may have differed in the preceding years. It is also relevant to point out, that the three months where the lockdown restrictions were most rigorous, might not necessarily be the peak months in terms of COVID-19 cases. Thus, the results from this study do not reflect the number of admissions during the months with most COVID-19 cases. Simple and subjective judgements were used of the level of lockdown restriction, whether the lockdown had an impact on the life of pregnant women and whether lockdown restrictions could possibly lead to non-admittance of EP infants. This may pose as a limitation in regards to response bias. It is plausible, however, that most of these biases would have induced a false association between the change in numbers and the possible explaining factors. Many of the principal investigators participating in this study, are representing NICUs from the same countries (e.g. five NICUs from Belgium). It is plausible that some EP infants are transferred between hospitals within the same country, which potentially could lead to a false high number of total admissions, since the same EP infant would count as an admission in more than one hospital. Such an issue would cause a false high number of admissions and thereby, a false high power. For EP infants re-admissions are likely, due to e.g. respiratory, gastro-intestinal or sensory problems. If re-admissions are included in the counted infants, this could also cause a false high power, however if it is done consistently for both 2019 and 2020 numbers, it will unlikely bias results. The analysis of number of admissions within each geographical region revealed no statistically significant differences between the three months where the lockdown restrictions were most rigorous in 2020 compared to the corresponding months of 2019. The p-values were adjusted to correct for multiple testing in the secondary outcome analyses. Furthermore, the small number of admissions which each region increases the risk of type II errors (10). Thus, studies with a larger number of admissions within each geographical region are needed.

The results from this study are contradictory to previously published literature (2)–(4) and may indicate, that when investigated on a global level, the effects of the lockdown caused by the COVID-19 pandemic on the rate of extremely preterm birth are small. Lockdown restrictions causing pregnant women to be more likely to stay home, may certainly influence several risk factors for preterm birth, such as stress and prolonged standing at work as well as infections on one side and the clinical processes leading to physician induced birth on the other side. Despite the number of EP admissions did not significantly differ between 2019 and 2020, the average number of randomisations dropped significantly in March 2020, where the COVID-19 pandemic spread across Europe. Possible explanations could be that several NICUs participating in the SafeBoosC-III trial, were forced to suspend all clinical research, due to the pandemic. In other NICUs, some staff members were re-allocated to clinical duty in COVID-19 departments. Furthermore, some NICUs split the clinical staff into smaller groups, so that fewer staff members were working simultaneously in the NICU, as a preventive measure to avoid the spread of COVID-19 within the department (personal communication). Such measures could all potentially affect the number of randomisations in SafeBoosC-III.

We have studied the immediate effects of lockdown. Delayed effects are also possible. The existence of neonatal networks and national registries will allow much stronger epidemiological studies to be done in the future, including examining the effects on the children themselves.

### Conclusion

This larger *ad hoc* study did not confirm previous studies’ report of a major reduction in the number of extremely preterm births during the first phase of the COVID-19 pandemic.

### What is already known about this topic

- Premature birth is the leading cause of under-five mortality globally
- Lockdown measures of the COVID-19 pandemic may have led to a decrease in the birth rate of extremely preterm infants

### What this study adds

- The number of extremely premature infant admissions to NICUs taking part in the international SafeBoosC-III trial did not significantly decrease during the three most rigorous months of lockdown restrictions caused by the first phase of the COVID-19 pandemic
- There were no differences in the change of extremely premature infant admissions across geographic regions during the COVID-19 lockdown
- Level of lockdown restrictions do not seem to be associated with a change in extremely preterm infant admissions

## Data Availability

The datasets used and/or analysed during the current study are available from the corresponding author on request and will be published on www.safeboosc.eu

## Acknowledgements

This study was conducted utilising the SafeBoosC-III consortium and we sincerely thank the principal investigators for their cooperation and rapid data collection.

## Funding

The sponsor/coordinating investigator, Professor of Paediatrics Gorm Greisen, is the initiator of the SafeBoosC-III project. He has no financial interest in the results of the trial. The Elsass Foundation, Aage and Johanne Louis-Hansen Foundation, and Svend Andersen Foundation supported the SafeBoosC-III trial through unconditional and unrestricted grants of DKK 2,700,000, DKK 1,000,000 and DKK 1,000,000, respectively. These funding sources had no role in the design of this study and will not have any role during its execution, analysis, interpretation of the data or decision to submit results.

## Consent for publication

Not applicable

## Competing interests

The authors declare that they have no competing interests.

## Author’s contributions

MIR, MLH and GG contributed to the conception and design of the study, drafted the protocol, collected data, drafted the manuscript, and will give final approval of the version to be published. TK, ED, AMH, SF, MLC, CL, ISK, GN, MW, EP, FM, RDR, JT, LC, LB, GH, PW, EM, JB, RV, DG, ZP, EH, ERW, HR, AEK, AM, SA, SP, BKO, KS, TS, OO, MS, AM, LA, EE, HF, JS, MB, AP, HO, LC, GP, MC, MA contributed to the data collection, revised the manuscript critically for important intellectual content, and will give final approval of the version to be published.

## Appendix 1

### Data sheet

1) **NICU:**
2) **Author and affiliation:**
3) **Start and end dates for three months of peak (defined as three consecutive months with most rigorous restrictions):**
4) **Extremely preterm infant count**

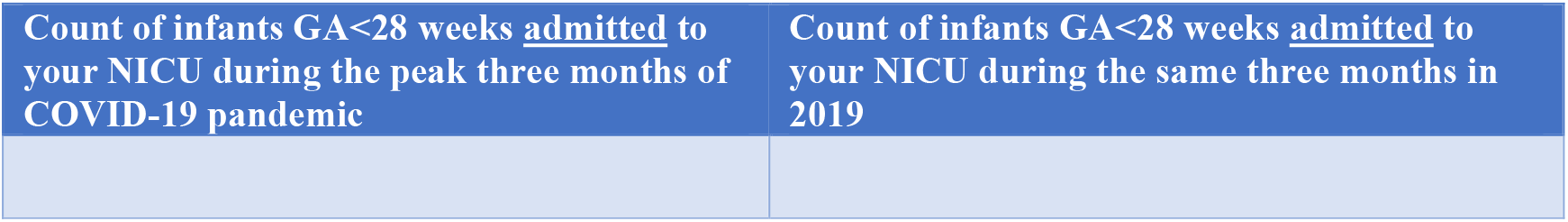
5) **Has there been any major changes in the organisation of perinatal care in your area which could be expected to change the number of admissions of extremely preterm infants to your NICU from 2019 to 2020? Tick the box with an x, which you find most correct**

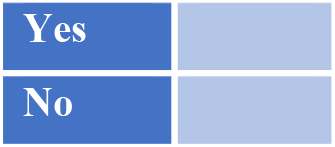 **If yes, please describe**:
6) **Please describe where/how you have obtained this data (admission logbook, NICU or hospital database, national registry, other: please describe):**
7) **On a scale from 1-5, one being very little change and five being radical change, how would you describe the impact of the COVID-19 lockdown on the everyday life of a pregnant woman (tick the box with an x, which you find most correct)**

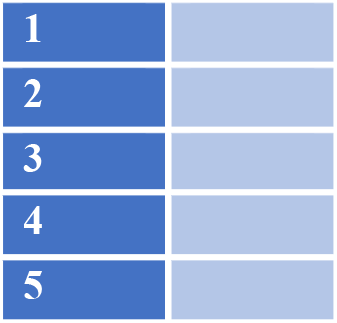
8) **On a scale from 1-5, one being very unlikely and 5 being almost certainly, how likely do you think that the COVID-19 restrictions outside and inside health institutions in your country/region has led to non-admittance of an extremely preterm infant. Causes could be intrauterine death, or no transfer from place of birth (tick the box with an x, which you find most correct)**

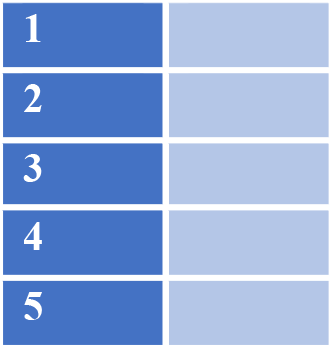
9) **On a scale from 1-5, one being no change in society and five being very strong change, would you describe the most rigorous restrictions in your country during the three peak months of the COVID-19 pandemic. Please base your answer on the following figure below:**

**Table 1:**
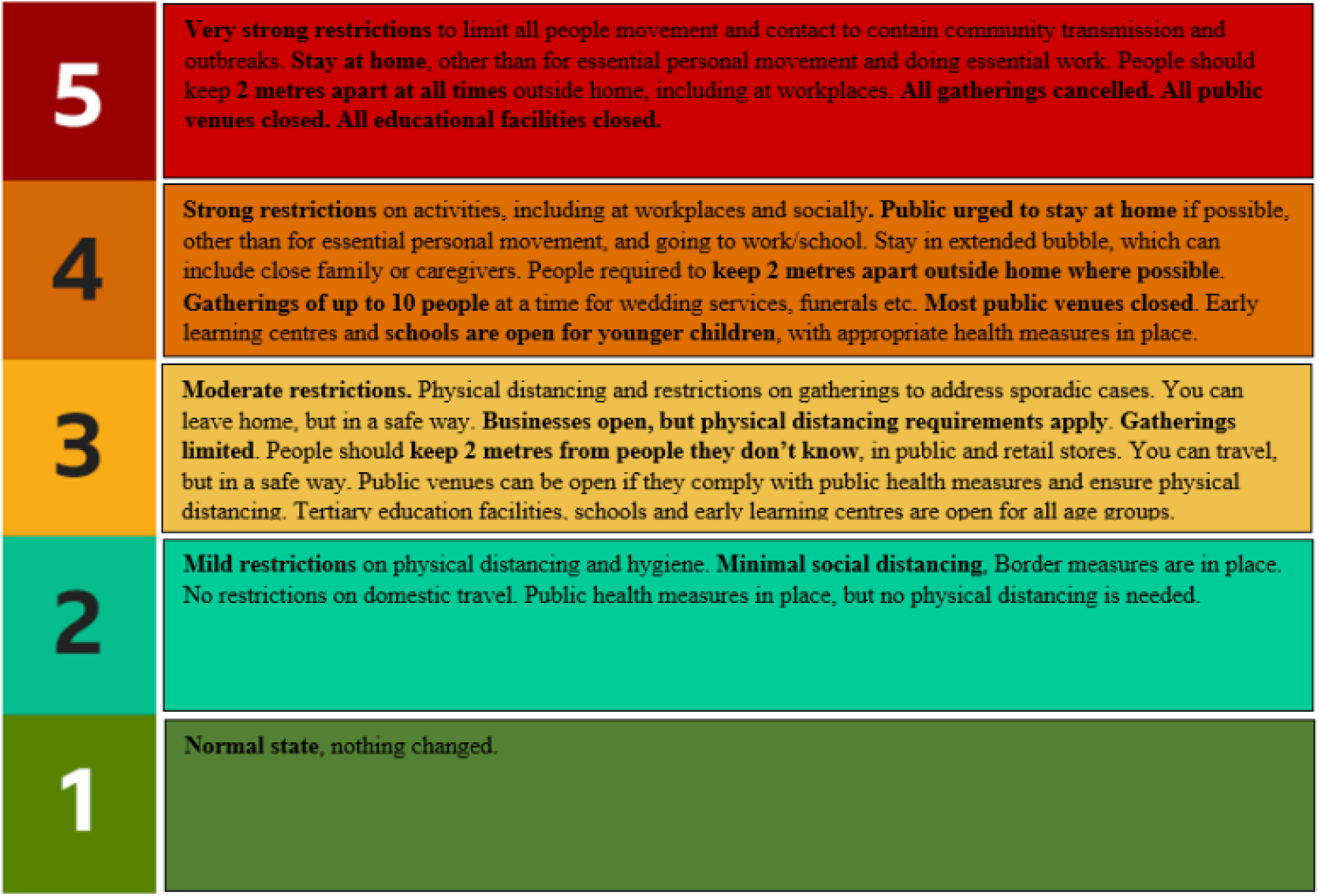
inspirations from the New Zealand COVID-19 alert system. **Tick the box with an x, which you find most correct:**

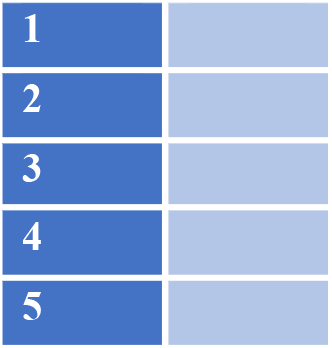

## Notes

### Competing Interest Statement

The authors have declared no competing interest.

### Clinical Trial

NCT04527601

### Clinical Protocols

https://clinicaltrials.gov/ct2/show/NCT04527601

### Author Declarations

According to Danish law, ethical permission to conduct this study was not necessary.

